# Comparative effects of CPAP and Mandibular Advancement Device Treatment on Cardiac Structure and Function in OSA: A Cardiovascular Magnetic Resonance Randomised Controlled Study

**DOI:** 10.1101/2025.02.12.25322131

**Authors:** Nithin R. Iyer, Yi-hui Ou, Jennifer A. Bryant, Thu-Thao Le, Juliana Tereza Colpani, Crystal S. Cheong, Weiqiang Loke, Peter A. Cistulli, Martin Ugander, Chi-hang Lee, Calvin W-L. Chin

**Affiliations:** Kolling Institute, Royal North Shore Hospital, and the University of Sydney, Sydney, Australia; National Heart Research Institute of Singapore, National Heart Centre Singapore, Singapore; Department of Medicine, Yong Loo Lin School of Medicine, National University of Singapore, Singapore; Cardiovascular Sciences ACP, Duke NUS Medical School, Singapore; Department of Endodontics, Operative Dentistry and Prosthodontics, Faculty of Dentistry, National University of Singapore, Singapore; Department of Otolaryngology–Head & Neck Surgery, National University Hospital, Singapore, Singapore; Sleep Research Group, Charles Perkins Centre and Northern Clinical School, Faculty of Medicine and Health, University of Sydney, Camperdown, New South Wales, Australia; Department of Respiratory and Sleep Medicine, Royal North Shore Hospital, St Leonards, New South Wales, Australia; Department of Clinical Physiology, Karolinska University Hospital, and Karolinska Institutet, Stockholm, Sweden; Cardiovascular Research Institute, National University Heart Centre, Singapore, Singapore; Department of Cardiology, National University Heart Centre, Singapore, Singapore; Department of Cardiology, National Heart Centre Singapore, Singapore

**Keywords:** Cardiovascular magnetic resonance, Continuous Positive Airway Pressure, Occlusal Splints, Hypertension, Sleep Apnea, Obstructive

## Abstract

**Aims:** Adverse cardiac remodelling is observed in obstructive sleep apnoea (OSA). However, the impact of OSA treatment on cardiac remodelling remains poorly defined. The Cardiosleep Research Program on Obstructive Sleep Apnoea, Blood Pressure Control and Maladaptive Myocardial Remodeling—Non-inferiority Trial (CRESCENT) showed that treatment with a mandibular advancement device (MAD) was non-inferior to continuous positive airway pressure (CPAP) for reducing 24-hour mean arterial BP in patients with hypertension and moderate-to-severe OSA. This cardiovascular magnetic resonance (CMR) substudy aimed to evaluate the comparative effects of MAD and CPAP on myocardial structure and function in patients with OSA.

**Methods and Results:** 85 patients with hypertension, increased cardiovascular risk and newly-diagnosed moderate-to-severe OSA (Apnoea-Hypopnoea Index [AHI] ≥15 events/hour), underwent contrast-enhanced CMR at baseline and following 12 months of treatment with CPAP (n=49) or MAD (n=36). LV mass, volumes, function and markers of diffuse myocardial fibrosis, including the extracellular volume (ECV) fraction and interstitial volume, were measured. Across the cohort, a small reduction in the ECV fraction was observed (25.0±2.0 vs 24.3±2.0%; p=0.002), accompanied by a reduction in the interstitial volume over a 12-month period of OSA treatment (24.0[21.3-27.7] vs 23.3[20.4-27.4]mL; p=0.044). The reduction in ECV fraction was similar between the CPAP and MAD groups (p=0.94).

**Conclusion:** 12 months treatment of OSA with either CPAP or MAD in patients with hypertension is associated with a small reduction in myocardial ECV fraction, a surrogate marker of diffuse myocardial fibrosis. Further confirmation of these findings in larger cohorts is warranted.

(Cardiosleep Research Program on Obstructive Sleep Apnea, Blood Pressure Control and Maladaptive Myocardial Remodeling—Non-inferiority Trial [CRESCENT]; NCT04119999).

## Introduction

Obstructive sleep apnoea (OSA) is a highly prevalent condition that is thought to affect nearly 1 billion people worldwide [1]. Patients with OSA develop recurrent collapse of the pharyngeal airway during sleep, resulting in intermittent hypoxaemia, sympathetic activation, and exaggerated intrathoracic pressure swings [2]. These perturbations are associated with acute and chronic haemodynamic effects, and particularly alterations in blood pressure [3, 4]. OSA is associated with several cardiovascular complications, including hypertension, atrial fibrillation, heart failure, coronary artery disease, stroke, pulmonary hypertension, metabolic syndrome, diabetes and sudden cardiac death.

Adverse cardiac remodelling is often observed in OSA, including left ventricular (LV) hypertrophy, left and right ventricular dysfunction, and focal myocardial fibrosis [5–7]. There is emerging evidence that treatment of OSA with continuous positive airway pressure (CPAP), the primary treatment for OSA, is associated with improvement in left ventricular function, in addition to right ventricular systolic function [8–10]. Mandibular advancement devices (MAD) treat OSA by advancing the mandible and are an effective alternative to CPAP, demonstrating improvement in OSA symptoms and quality of life [11–13]. Few studies have assessed the effect of MAD on ventricular structure and function in patients with OSA. One echocardiographic study showed improvement in LV hypertrophy in patients with OSA treated with MAD for six months [14]. Additionally, it is not known whether treatment of OSA, with either MAD or CPAP, can reverse myocardial fibrosis.

Cardiovascular magnetic resonance (CMR) imaging has become the non-invasive reference standard for interrogating the myocardium due to its ability to accurately assess cardiac morphology, function, and myocardial tissue characteristics. In particular, late gadolinium enhancement (LGE) permits visualisation of focal replacement myocardial fibrosis, while T1 mapping pre- and post-gadolinium contrast enables non-invasive measurement of the myocardial extracellular volume (ECV) fraction, a quantitative marker of myocardial diffuse interstitial fibrosis.

Recently, the CRESCENT randomised trial (Cardiosleep Research Program on Obstructive Sleep Apnoea, Blood Pressure Control and Maladaptive Myocardial Remodeling—Non-inferiority Trial) showed that MAD was non-inferior to CPAP for reducing 24-hour mean arterial BP in patients with hypertension and moderate-to-severe OSA, after 6 months of treatment [13]. The present study was a cardiac MRI substudy of the CRESCENT trial and was designed to determine the effects of MAD and CPAP on myocardial structure and function, including markers of myocardial fibrosis, in patients with moderate-to-severe OSA.

## Methods

### Study Population and Trial Design

The CRESCENT trial design has been described previously [13]. Briefly, this was a randomized, controlled, non-inferiority trial conducted in Singapore (NCT04119999). Participants were recruited from October 1, 2019, to December 5, 2022, through medical record screening at three public hospitals in Singapore. Inclusion criteria consisted of adults aged ≥40 years with known hypertension and at least one other factor for high cardiovascular risk for screening polysomnography, including diabetes mellitus; stroke or transient ischaemic attack; significant coronary artery disease (at least one stenosis of >50% in at least one major epicardial artery on invasive or CT coronary angiography, previous coronary revascularisation, previous myocardial infarction or abnormal stress test); chronic kidney disease with an estimated glomerular filtration rate of less than 60mL/min/1.73m^2^ or age of 75 years or older [13]. Exclusion criteria included known OSA on treatment; Cheyne-Stokes breathing or predominantly central sleep apnoea; secondary hypertension; unsuitable anatomy for MAD; life expectancy <1 year; hypertensive crisis, acute coronary syndromes, or acute heart failure in the past 30 days.

Participants underwent polysomnography following recruitment, and completed the Epworth Sleepiness Scale (ESS) questionnaire. OSA was diagnosed based on the apnoea-hypopnoea index (AHI), quantified as the total number of apnoeas or hypopnoeas per hour of sleep. Patients diagnosed with moderate-to-severe OSA (AHI ≥15 events per hour) were randomly assigned to treatment by MAD or CPAP in a 1:1 ratio. The treatment duration was 12 months. Device adherence was determined in the MAD group by an embedded compliance micro-recorder chip (DentiTrac, Braebon), and in the CPAP group by a cloud-based telemedicine management platform (Airview, ResMed Corp). Device adherence was defined as ≥4 hours usage per night, beyond which CPAP treatment may reduce the incidence of cardiovascular events [15]. The Institutional Review Board approved the trial (The Domain Specific Review Board-C: 2019/00359, approved on August 28, 2019). All participants provided written informed consent.

### Cardiovascular magnetic resonance image acquisition

All patients in the CRESCENT cohort were invited and assessed for suitability for CMR, up to a pre-specified total of 100 patients. Those who agreed and were eligible underwent a standardised CMR protocol with a Siemens Aera 1.5 Tesla MRI scanner (Siemens Healthineers, Erlangen, Germany) at baseline and following 12 months of treatment with MAD or CPAP. Balanced steady-state free-precession cines were acquired in the standard long-axis views and a short-axis stack was acquired from base to apex. LGE images were acquired at 10 min after 0.1 mmol/kg of gadobutrol (Gadovist®, Bayer Pharma AG, Germany) with a phase-sensitive inversion-recovery gradient-echo imaging sequence. The inversion time for optimal myocardial nulling was selected from an inversion time scout sequence. T1 maps were acquired at the basal and mid-ventricular short-axis levels, pre- and 15-min post-contrast with modified Look-Locker inversion-recovery (MOLLI) 5s(3s)3s and 4s(1s)3s(1s)2s acquisition schemes, respectively.

### CMR analysis

Image analysis was performed using CVI42 software (Circle Cardiovascular Imaging, Calgary, Canada) by trained imaging fellows at the National Heart Research Institute of Singapore CMR Core Laboratory, who were blinded to the patients’ clinical status. Ventricular volumes, mass and ejection fraction, were measured from the short-axis cine stack, using manual contouring of the left ventricle in end-diastole and end-systole, excluding papillary muscles, as detailed previously [16]. LV volumes and mass were indexed to body surface area. The presence of LGE was assessed qualitatively by two readers according to the recommendations by the Society of CMR [17]. Average native and post-contrast myocardial T1 values were measured by placing a region of interest (ROI) within the middle third of the short-axis myocardial wall at the basal- and mid-ventricular levels while avoiding areas of focal LGE. The myocardium-blood pool interface was carefully avoided to prevent partial volume effects. Pre- and post-contrast blood T1 values were measured in a ROI drawn within the blood pool. The ECV fraction was calculated from the pre-and post-contrast average blood and myocardial T1 values, as described previously [18, 19], and is reported as a mean value from the basal and mid-ventricular slices. The myocardial interstitial volume was calculated as: 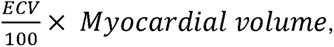 where myocardial volume (mL) was defined as myocardial mass (g)/1.05 g/mL. Myocyte volume was calculated as: 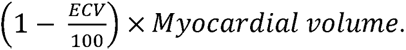 Myocardial strain was analysed in the cine images using the Tissue Tracking Plugin (Circle Cardiovascular Imaging, Calgary, Canada).

### Statistics

Normality for continuous variables was assessed using the Shapiro-Wilk test. Normally distributed data are presented as mean ± standard deviation. Non-normally distributed data are presented as median [interquartile range]. Comparisons were performed for continuous variables before and after treatment using the parametric paired Student *t*-test or the non-parametric Wilcoxon signed-ranked test. Pearson’s correlation coefficient was used to assess correlations between continuous variables. Categorical variables are presented as number (percentage) and compared using the χ^2^ test. Statistical analyses were performed using SPSS Version 29 (Statistical Package for the Social Sciences, International Business Machines, Inc., Armonk, New York, USA) and GraphPad Prism 9.4.1 (GraphPad Software, Inc., San Diego, California, USA). A two-sided p-value <0.05 was considered as statistically significant.

## Results

Of the 220 participants in the CRESCENT study who were randomly allocated to either MAD or CPAP in a 1:1 allocation, 101 patients agreed and were eligible to participate in the CMR substudy. These patients underwent baseline CMR (Figure 1). Out of this group, 84.2% (85 of 101) underwent serial CMR at 12 months and were included in this sub-study (median age: 61 years; 91% [77 of 85] males; MAD: 36; CPAP: 49).

**Figure 1.**
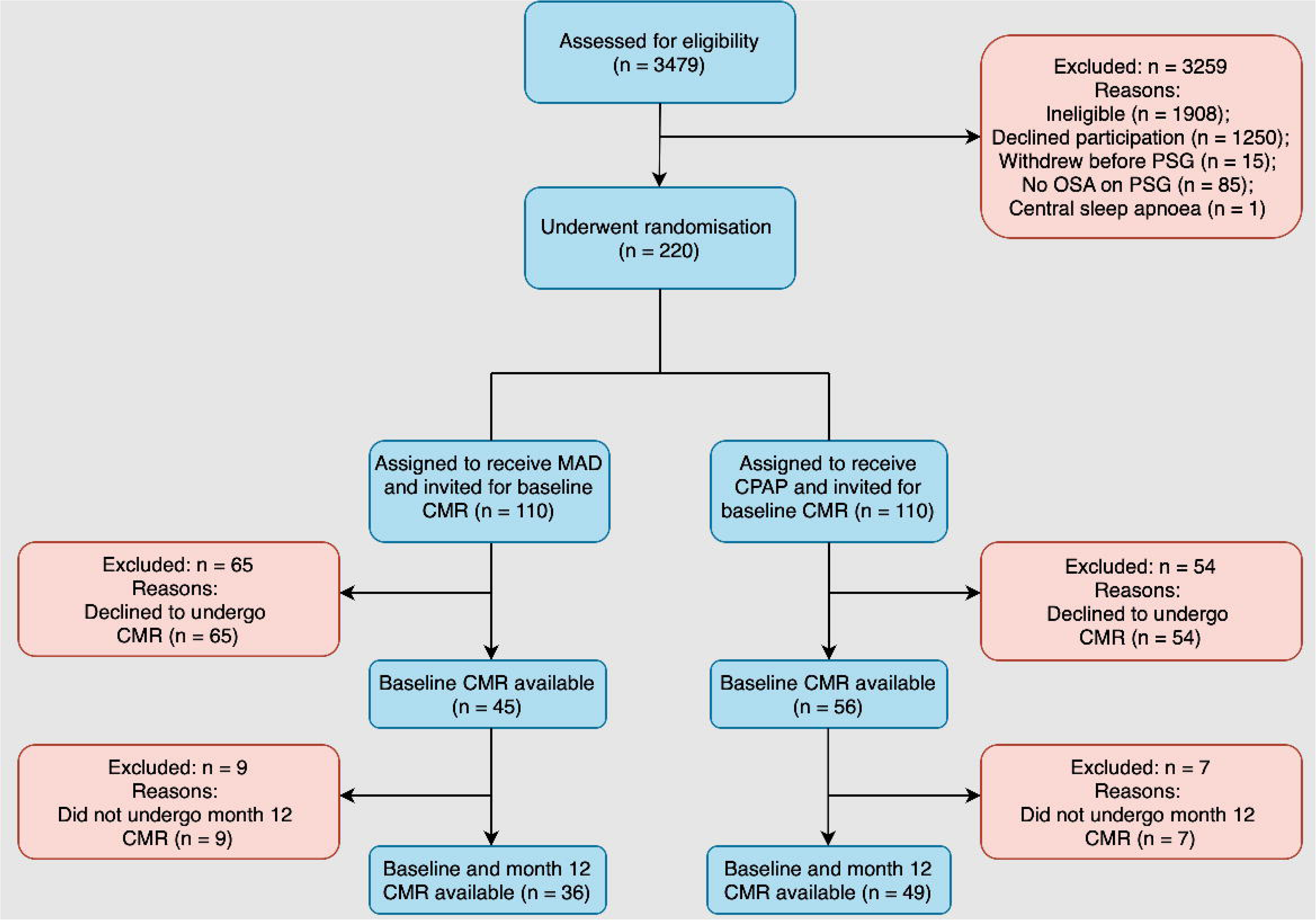
Flow chart of patient inclusion. Abbreviations: OSA: Obstructive sleep apnoea; PSG: polysomnography; MAD: Mandibular advancement device; CPAP: Continuous positive airways pressure; CMR: Cardiovascular magnetic resonance.

At baseline, the median BMI was 28 [26-31] kg/m^2^. The baseline 24-hour mean arterial BP was 96±8 mmHg. Of the cardiovascular co-morbidities, 61% (52 of 85) had coronary artery disease, and 60% (51 of 85) had diabetes mellitus. All patients had hypertension. OSA was classified as severe in 68% (58 of 85) of patients. The baseline median AHI for the cohort was 41 [27-59] events per hour. The ESS at baseline was 7.8 ± 4.3. The baseline CMR characteristics are given in Table 1. Patients in the MAD group were older compared to the CPAP group (MAD: 64 [57-69] years vs CPAP: 59 [53-64] years; p = 0.043). Otherwise, there were no clinical, sleep or CMR differences in baseline characteristics between the MAD and CPAP groups (Supplemental Table 1).

**Table 1.**
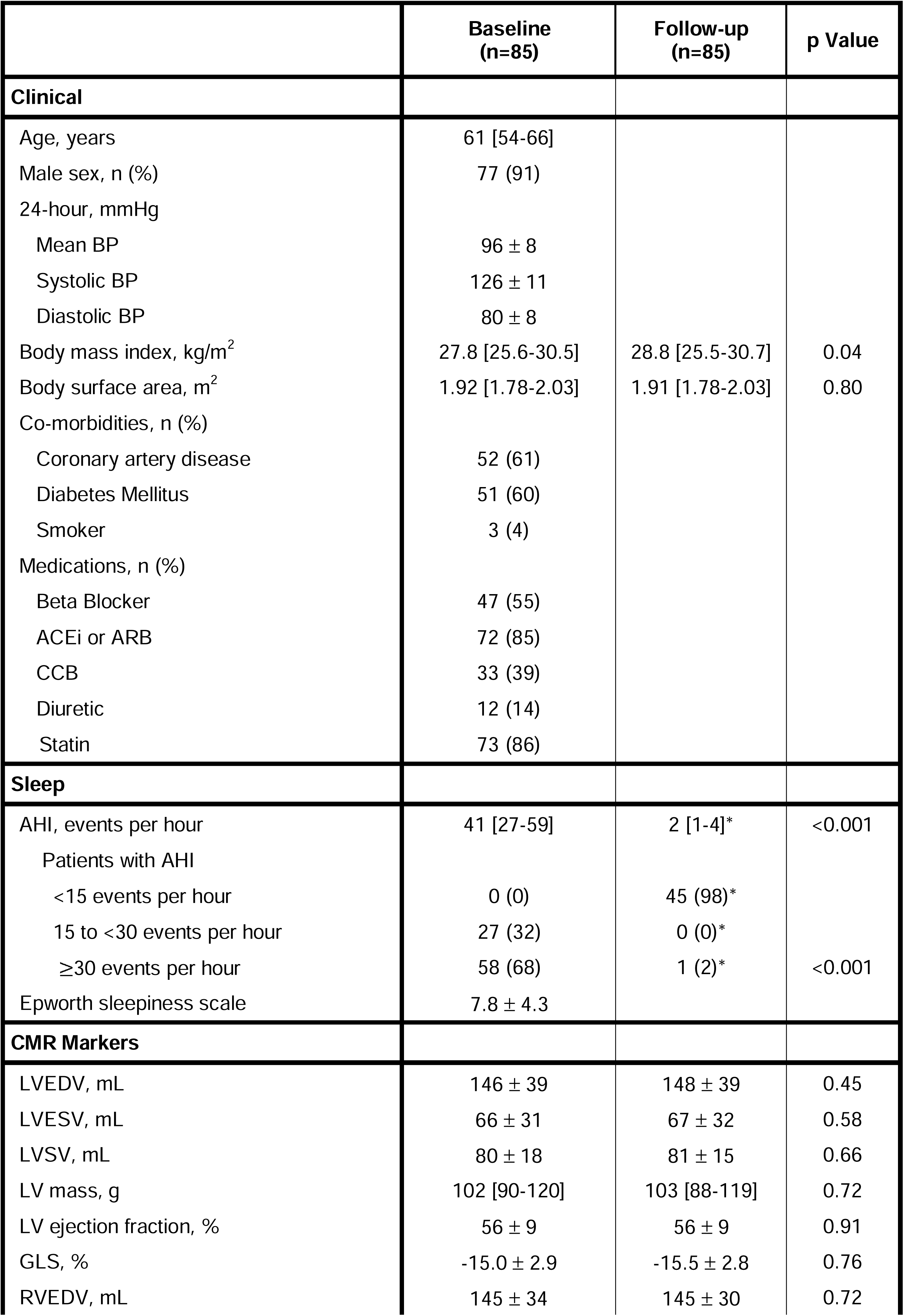
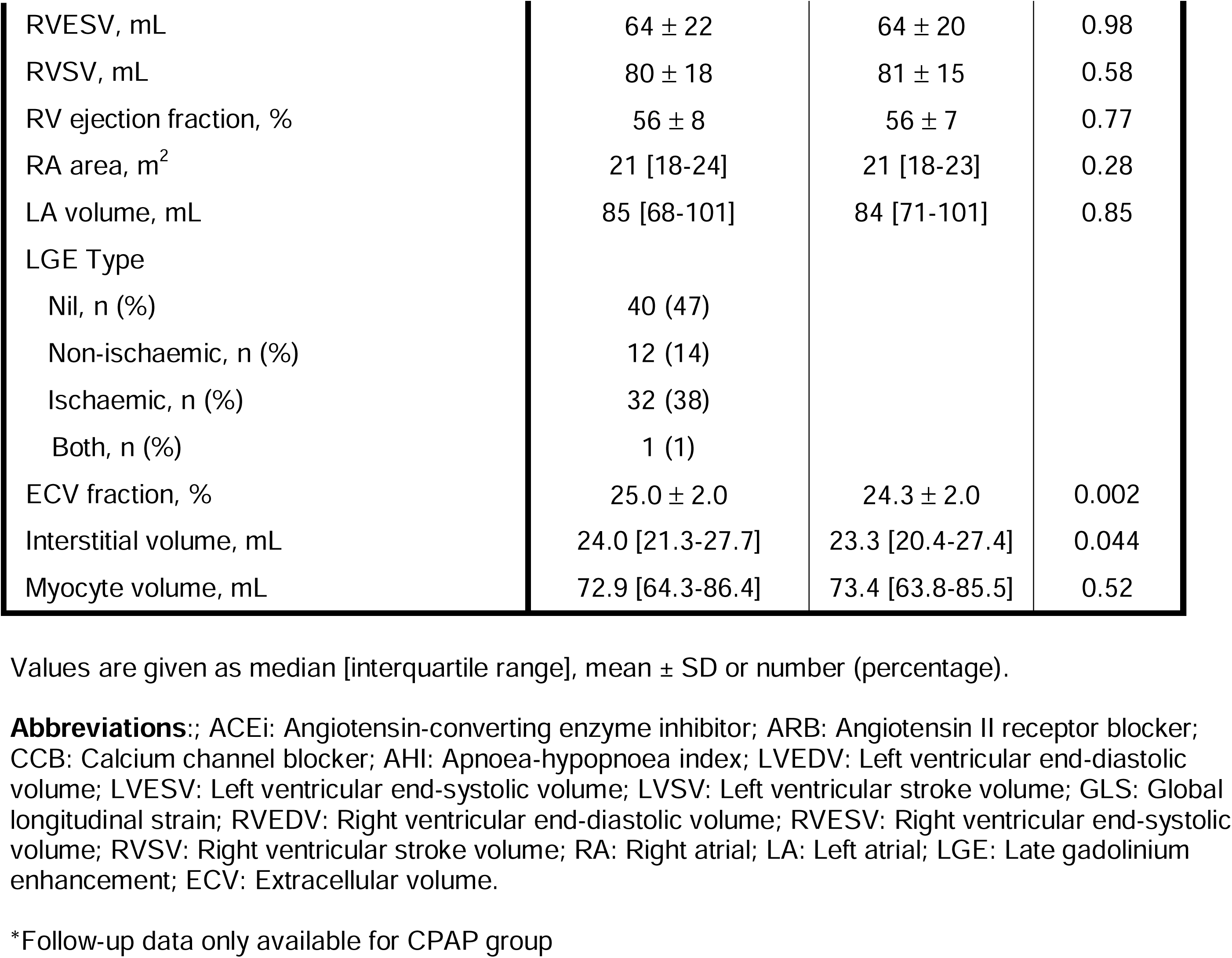
Clinical, sleep and CMR characteristics of the cohort at baseline and following 12 months of OSA treatment.

Valid adherence data at 12 months was available for 58% (21 of 36) in the MAD group. The mean duration of device usage per night was 4.6±2.4 hours in this group. In the MAD group, 62% (13 of 21) used the device for ≥4 hours per night and were classified as adherent. Adherence data was available for 92% (45 of 49) in the CPAP group. The mean duration of device usage per night in this group was 4.3±2.0 hours, with 64% (29 of 45) being adherent. The difference between groups in adherence was not statistically significant (p=0.57).

In the MAD group, the ECV fraction decreased from 24.9±2.3% at baseline to 24.0±2.2% at month 12; p=0.042). The ECV fraction decreased in the CPAP group from 25.1±1.9% at baseline to 24.5±1.8% at month 12 (p=0.016). Change in ECV fraction did not differ according to treatment device (MAD: -0.6[-1.6-0.4] % vs CPAP: -0.6[-2.1-0.5] %; p=0.94). Amongst the CMR markers in the entire cohort, there was a reduction in the ECV fraction (baseline: 25.0±2.0 vs month 12: 24.3±2.0%; p=0.002), with associated reduction in the interstitial volume (24.0[21.3-27.7] vs 23.3[20.4-27.4] mL; p=0.044) (Figure 2). All other CMR parameters in the entire cohort, including LV mass and LV myocyte volume, did not differ, and these results were consistent across treatment groups (Table 2).

**Figure 2.**
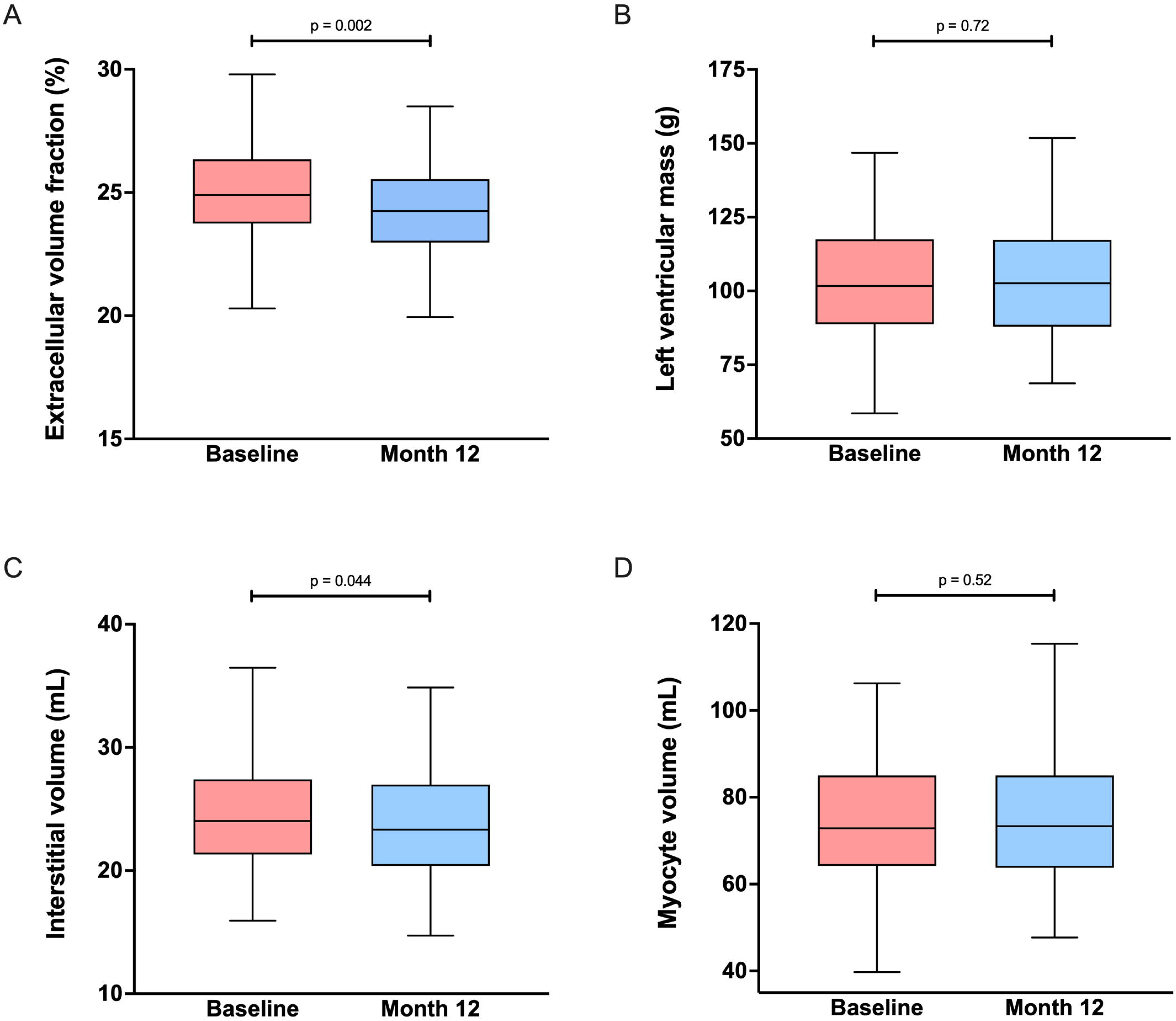
Extracellular volume fraction (ECV) fraction before and after OSA treatment. ECV fraction (panel A) and absolute myocardial interstitial volume (panel C) were reduced following 12 months of treatment for OSA. Left ventricular mass (panel B) and myocyte volume (panel D) did not change following treatment. Whiskers were plotted using the Tukey method.

**Table 2.**
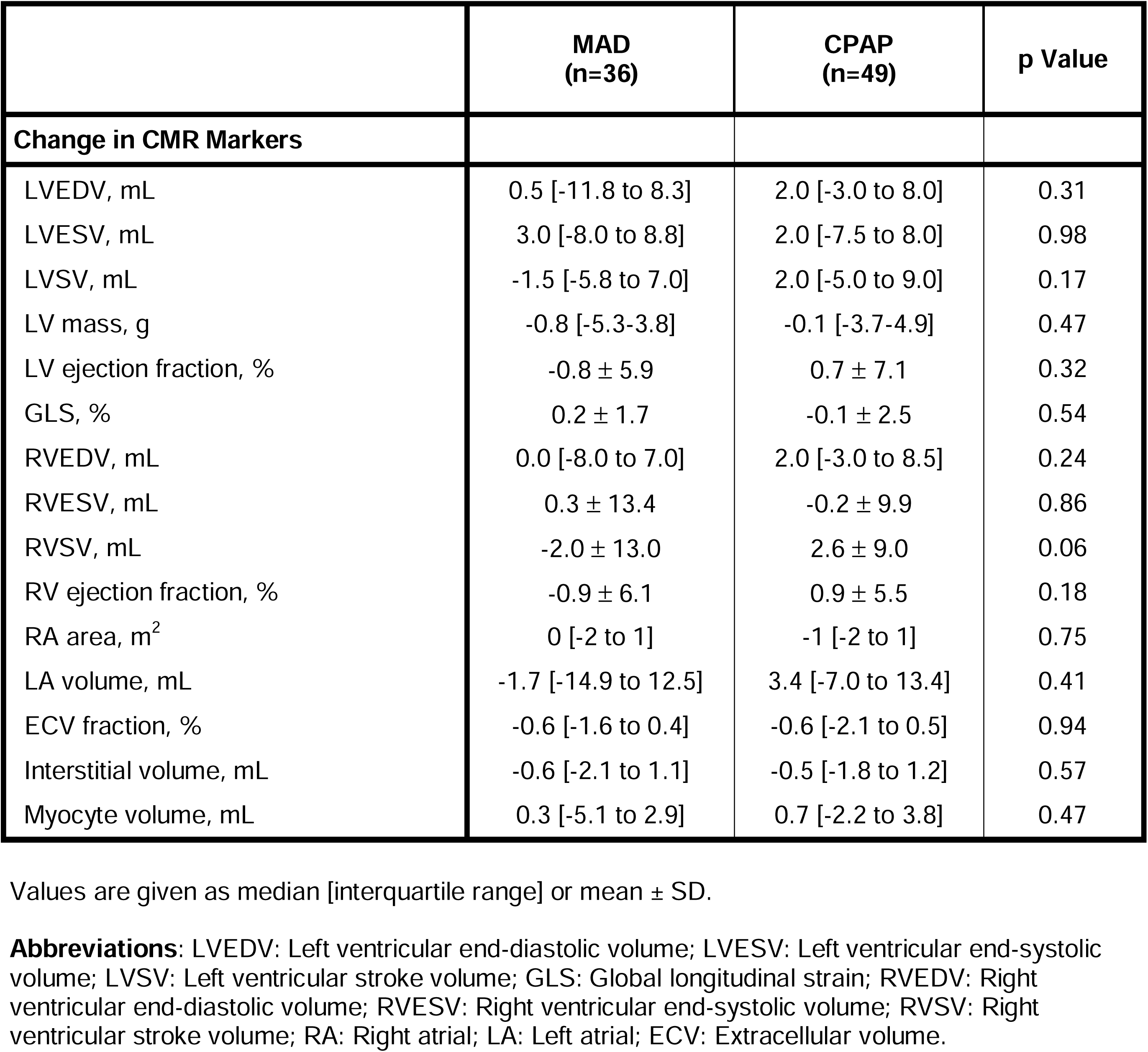
Change in CMR characteristics of the cohort following 12 months of treatment for OSA according to treatment group.

|  | MAD<br>(n=36) | CPAP<br>(n=49) | p Value |
| --- | --- | --- | --- |
| **Change in CMR Markers** |  |  |  |
| LVEDV, mL | 0.5 [-11.8 to 8.3] | 2.0 [-3.0 to 8.0] | 0.31 |
| LVESV, mL | 3.0 [-8.0 to 8.8] | 2.0 [-7.5 to 8.0] | 0.98 |
| LVSV, mL | -1.5 [-5.8 to 7.0] | 2.0 [-5.0 to 9.0] | 0.17 |
| LV mass, g | -0.8 [-5.3-3.8] | -0.1 [-3.7-4.9] | 0.47 |
| LV ejection fraction, % | -0.8 ± 5.9 | 0.7 ± 7.1 | 0.32 |
| GLS, % | 0.2 ± 1.7 | -0.1 ± 2.5 | 0.54 |
| RVEDV, mL | 0.0 [-8.0 to 7.0] | 2.0 [-3.0 to 8.5] | 0.24 |
| RVESV, mL | 0.3 ± 13.4 | -0.2 ± 9.9 | 0.86 |
| RVSV, mL | -2.0 ± 13.0 | 2.6 ± 9.0 | 0.06 |
| RV ejection fraction, % | -0.9 ± 6.1 | 0.9 ± 5.5 | 0.18 |
| RA area, m<sup>2</sup> | 0 [-2 to 1] | -1 [-2 to 1] | 0.75 |
| LA volume, mL | -1.7 [-14.9 to 12.5] | 3.4 [-7.0 to 13.4] | 0.41 |
| ECV fraction, % | -0.6 [-1.6 to 0.4] | -0.6 [-2.1 to 0.5] | 0.94 |
| Interstitial volume, mL | -0.6 [-2.1 to 1.1] | -0.5 [-1.8 to 1.2] | 0.57 |
| Myocyte volume, mL | 0.3 [-5.1 to 2.9] | 0.7 [-2.2 to 3.8] | 0.47 |

Values are given as median [interquartile range] or mean ± SD.
**Abbreviations:** LVEDV: Left ventricular end-diastolic volume; LVESV: Left ventricular end-systolic volume; LVSV: Left ventricular stroke volume; GLS: Global longitudinal strain; RVEDV: Right ventricular end-diastolic volume; RVESV: Right ventricular end-systolic volume; RVSV: Right ventricular stroke volume; RA: Right atrial; LA: Left atrial; ECV: Extracellular volume.

An exploratory analysis of clinical and CMR markers associated with ECV fraction change showed a reduction in the ECV fraction in obese patients (baseline BMI ≥30kg/m^2^; baseline ECV: 25.2[23.9-26.6]% vs month 12 ECV: 23.9[22.5-25.3]%; n=25; p<0.001). Furthermore, obese patients were more likely to experience a reduction in the ECV fraction (change in obese: -1.2[-1.9 to -0.3]% vs change in non-obese: -0.4[-1.6 to 0.8]%; p=0.026) (Figure 3). Obese patients had a higher AHI at baseline (54[42-69] events per hour vs 32[23-48] events per hour; p<0.001), and experienced a greater reduction in AHI with treatment (58[42-66] events per hour vs 31[23-46] events per hour; p=0.001).

**Figure 3.**
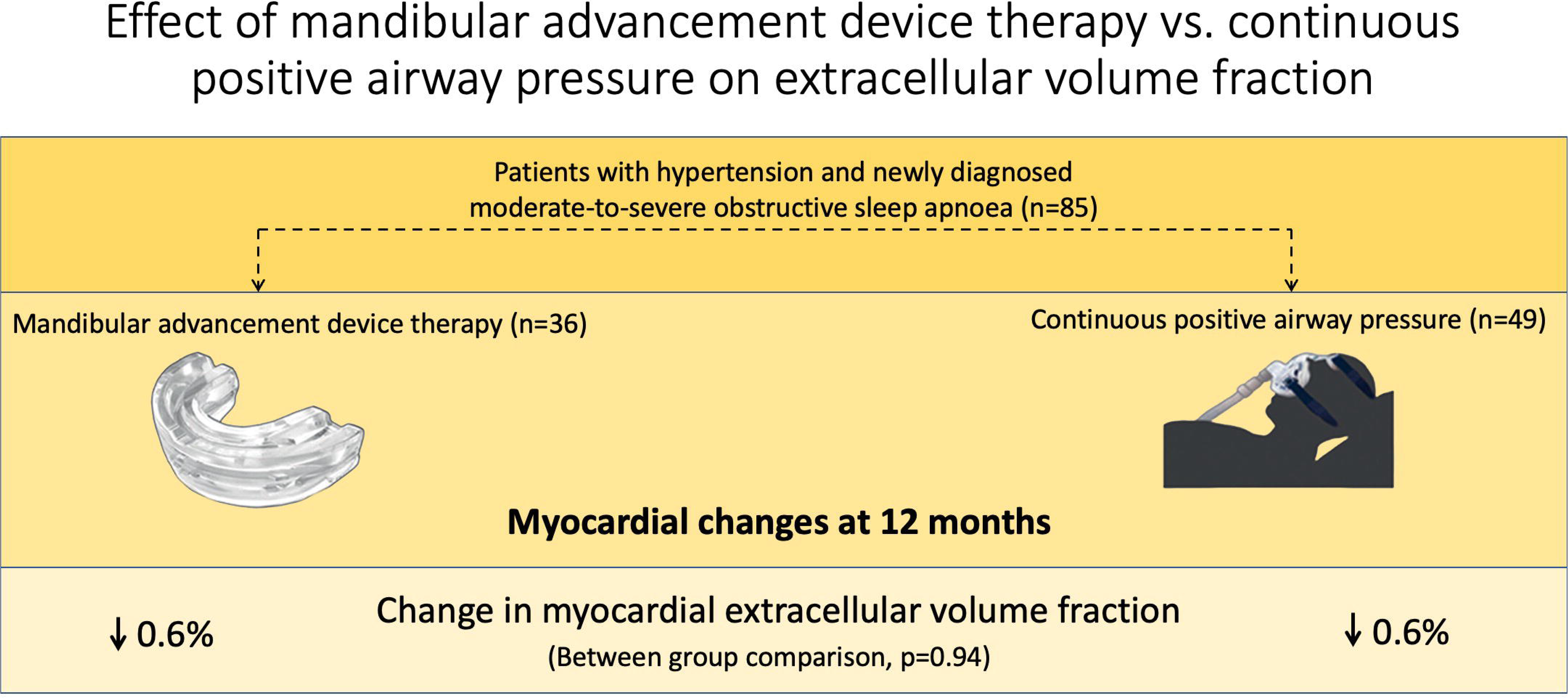
Forest plot for subgroup analyses of treatment effect. Patients with obesity (BMI ≥30kg/m^2^) were more likely to experience a reduction in ECV fraction with OSA treatment. Median and interquartile range are shown for all subgroup analyses. Dotted line shows no effect point. Abbreviations: BMI: Body mass index; SBP: Systolic blood pressure; AHI: Apnoea-hypopnoea index; ACEi: Angiotensin-converting enzyme inhibitor; ARB: Angiotensin II receptor blocker.

Otherwise, there were no associations detected between ECV fraction change and change in clinical, sleep or other CMR markers (Supplemental Table 2).

## Discussion

To our knowledge, this is the first CMR study to assess the comparative effects of MAD and CPAP therapy for OSA on longitudinal change in cardiac functional parameters. The main finding in this study was that treatment of OSA, with either MAD or CPAP, was associated with a small reduction in the ECV fraction, a surrogate marker of diffuse myocardial fibrosis. Conversely, no change was observed in LV mass or function.

OSA is thought to mediate myocardial fibrosis via several mechanisms. Chronic intermittent hypoxia, a hallmark of OSA, has been shown to induce cardiac inflammation, apoptosis and fibrosis in rats [20]. Heightened sympathetic drive due to recurrent apnoeas during sleep has been implicated in the development of cardiac interstitial fibrosis in animal models [21, 22]. Resting awake OSA patients have increased BP variability, which has been shown to induce myocardial fibrosis in hypertensive rats [22–24]. Alternating cycles of hypoxaemia and reoxygenation may trigger oxidative stress mechanisms, which are known to be involved in the development of myocardial fibrosis [22, 25]. Finally, hypertension, commonly found in patients with OSA, is known to cause both replacement and diffuse myocardial fibrosis [26, 27].

Treatment of OSA is known to target a number of pathways that have been linked to the development of myocardial fibrosis. The evidence is strongest for CPAP. In particular, CPAP has been shown to reduce sympathetic drive and acutely decrease BP during sleep [28, 29]. The chronic effects of CPAP on BP are more modest, with greater benefit evident in hypertensive rather than normotensive patients [30]. CPAP reduces nocturnal, in addition to waking, BP variability [31, 32]. Markers of oxidative stress are reduced with CPAP therapy [33, 34]. CPAP has also been shown to improve nocturnal hypoxemia [35]. Therefore, it is plausible that CPAP may reduce myocardial fibrosis in OSA through these mechanisms.

Myocardial fibrosis includes both replacement fibrosis and reactive interstitial fibrosis. Replacement fibrosis, represented by late gadolinium enhancement in CMR imaging, corresponds to the replacement of myocytes with collagen following myocyte necrosis and is thought to be irreversible [36]. Reactive interstitial fibrosis, measured with T1 mapping and ECV fraction at CMR, is characterised by collagen deposition by myofibroblasts within the interstitium and perivascular space. It is considered a marker of intermediate disease severity and potentially reversible [37, 38]. The observed reduction in the ECV fraction, along with a reduction in the absolute myocardial interstitial volume, indicates that treatment of OSA with MAD or CPAP may specifically reduce the burden of myocardial fibrosis through effects on the interstitium. However, it must be noted that most patients in this cohort had normal ECV fraction values at baseline [39] and the absolute reduction in ECV fraction was small, within the expected measurement variability. Therefore, the observed change in ECV fraction is unlikely to be clinically significant in this cohort. Future work should evaluate whether a cohort with more advanced OSA (with poorer control of hypertension, higher rates of obesity and/or heart failure with preserved ejection fraction), and therefore with higher expected baseline myocardial fibrosis [40–42], might derive more clinically meaningful improvement in ECV fraction with OSA treatment.

Myocardial fibrosis regression has been documented previously following interventions in other cardiac diseases, including aortic valve replacement for aortic stenosis and pirfenidone, an oral antifibrotic agent, for heart failure with preserved ejection fraction [38, 43]. Both studies observed changes in ECV fraction and interstitial volume after one year of treatment. The magnitude of ECV fraction reduction was higher with pirfenidone (-1.21% compared to placebo) compared to this study. This likely reflects the more advanced heart failure population in that study, with baseline mean myocardial ECV fraction measuring 30.1%. ECV fraction was paradoxically increased following aortic valve replacement for aortic stenosis due to greater effects on the myocyte compartment.

The reduction in ECV fraction was noted across both MAD and CPAP groups. A growing body of evidence suggests that MAD may have similar cardiovascular benefits to CPAP. MAD has been shown to improve nocturnal hypoxaemia [44]. MAD has a similar BP-lowering effect compared to CPAP [13]. There is emerging evidence to suggest that MAD treatment can improve cardiac autonomic function [45]. Based on these benefits, it is plausible that MAD therapy would also be able to reduce myocardial fibrosis. This is an important finding, since MADs are often preferred by patients with OSA compared to CPAP [46], indicating great potential to improve cardiovascular health.

In this study, there was no difference observed in left ventricular mass, atrial and ventricular volumes, and ventricular function following treatment for OSA. Prior to this study, a few longitudinal CMR studies have assessed for change in cardiac functional parameters following initiation of CPAP in OSA. Results have been mixed. One study showed reduction in indexed right and left atrial volumes, indexed RV end diastolic volume, and indexed left ventricular mass at 1 year following initiation of CPAP therapy [8]. Notably, the cohort in that study at baseline was generally heavier (mean BMI 38 kg/m^2^), more symptomatic (mean ESS 14) and had more severe OSA (mean AHI 63 events/hour), which may have explained the discrepancy in findings compared to our study. Two studies have shown improvement in RV volumes or RV ejection fraction with CPAP therapy [9, 47]. In both studies, patients had a higher BMI at baseline (33-35 kg/m^2^) compared to our study (28 kg/m^2^). Another study showed no change in CMR parameters after four months of CPAP therapy in non-obese and obese CPAP-naïve patients [48]. The patients included were most similar to our cohort based on baseline symptoms (mean ESS 9.5), BMI (31.6 kg/m^2^) and severity of OSA (mean AHI 36 events/hour), which may have explained the similarity in findings.

The finding of a change in myocardial fibrosis markers without associated changes in cardiac mass, volumes or function has precedent. Recently, pirfenidone was found to reduce the ECV fraction in patients with heart failure without any effects on cardiac functional parameters or LV mass [38]. This is an antifibrotic agent without haemodynamic effect and is therefore thought to target the extracellular matrix. In an earlier study, lisinopril caused regression of myocardial fibrosis in patients with hypertensive heart disease, as measured with the LV collagen fraction and myocardial hydroxyproline concentration from endomyocardial biopsy, without any effects on LV hypertrophy [49]. These results were in keeping with the perception that haemodynamics are the most relevant determinants of LVH. Since the BP was controlled at baseline, no changes in LVH were detected [49]. Similarly, the patients in our study had well-controlled BP at baseline, which may have explained why no regression in LV mass was observed.

### Limitations

Our study has a few limitations. First, this was an exploratory analysis in a CMR substudy of the CRESCENT trial. The hypothesis that OSA treatment would reduce the myocardial ECV fraction was not *a priori*. Second, this was a single-centre CMR study. Although the sample size was small, this was similar to previous CMR studies assessing the longitudinal effect of OSA treatment on cardiac functional parameters. Third, the participants were all of Chinese ethnicity and mostly male (91%), which may limit the generalisability [13].

## Conclusion

Treatment of moderate-to-severe OSA, with either MAD or CPAP, in patients with hypertension and high cardiovascular risk, is associated with a small reduction in myocardial ECV fraction, a surrogate marker of diffuse myocardial fibrosis, at 12 months, with an associated reduction in the myocardial interstitial volume.

## Data Availability

The datasets generated and analysed for the current study are not publicly available. All data produced in the present study are available upon reasonable request to the authors.

## Data Availability Statement

The datasets generated and analysed for the current study are not publicly available. Please contact the corresponding author for data requests.

## Non-standard Abbreviations and Acronyms

CMR: Cardiovascular magnetic resonance
CPAP: Continuous positive airway pressure
ECV: Myocardial extracellular volume fraction
LV: left ventricle
MAD: Mandibular advancement device
OSA: Obstructive sleep apnoea

