## Supplemental Tables for "Comparative effects of CPAP and Mandibular Advancement Device Treatment on Cardiac Structure and Function in OSA: A Cardiovascular Magnetic Resonance Randomised Controlled Study"

**Supplemental Table 1. Clinical, sleep and CMR characteristics of the cohort at baseline according to treatment group.**

|  | **MAD**  **(n=36)** | **CPAP**  **(n=49)** | **p Value** |
| --- | --- | --- | --- |
| **Clinical** |  |  |  |
| Age, years | 64 [57-69] | 59 [53-64] | 0.04 |
| Male sex, n (%) | 34 (94) | 43 (88) | 0.30 |
| 24-hour, mmHg |  |  |  |
| Mean BP | 95 ± 9 | 96 ± 7 | 0.32 |
| Systolic BP | 125 ± 11 | 127 ± 10 | 0.34 |
| Diastolic BP | 80 ± 9 | 81 ± 7 | 0.38 |
| Body mass index, kg/m^2^ | 27.5 [25.3-29.9] | 28.3 [25.8-30.8] | 0.45 |
| Body surface area, m^2^ | 1.95 [1.78-2.10] | 1.91 [1.76-2.01] | 0.36 |
| Co-morbidities, n (%) |  |  |  |
| Coronary artery disease | 24 (67) | 28 (57) | 0.37 |
| Diabetes Mellitus | 22 (61) | 29 (59) | 0.86 |
| Smoker | 2 (6) | 1 (2) | 0.39 |
| Hypertension duration, y |  |  | 0.24 |
| <5, n (%) | 5 (14) | 13 (27) |  |
| 5 – 10, n (%) | 7 (19) | 7 (14) |  |
| >10, n (%) | 11 (31) | 19 (39) |  |
| Unknown, n (%) | 13 (36) | 10 (20) |  |
| Medications, n (%) |  |  |  |
| Beta Blocker | 20 (56) | 27 (55) | 0.97 |
| ACEi or ARB | 29 (81) | 43 (88) | 0.36 |
| CCB | 15 (42) | 18 (37) | 0.65 |
| Diuretic | 5 (14) | 7 (14) | 0.96 |
| Statin | 33 (92) | 40 (82) | 0.19 |
| **Sleep** |  |  |  |
| AHI, events per hour | 33 [22-54] | 41 [26-58] | 0.29 |
| Patients with AHI |  |  | 0.46 |
| 15 to <30 events per hour | 13 (36) | 14 (29) |  |
| $\geq$30 events per hour | 23 (64) | 35 (71) |  |
| Epworth sleepiness scale | 7.6 ± 4.7 | 8.0 ± 3.9 | 0.66 |
| **CMR Markers** |  |  |  |
| LVEDV, mL | 147 [115-173] | 139 [120-164] | 0.70 |
| LVESV, mL | 61 [44-85] | 59 [50-73] | 0.98 |
| LVSV, mL | 80 ± 17 | 80 ± 19 | 0.92 |
| LV mass, g | 104 [89-123] | 102 [90-114] | 0.40 |
| LV ejection fraction, % | 58 [50-64] | 57 [52-62] | 0.63 |
| GLS, % | -14.9 ± 3.1 | -15.0 ± 2.8 | 0.78 |
| RVEDV, mL | 146 ± 38 | 144 ± 31 | 0.84 |
| RVESV, mL | 65 ± 25 | 64 ± 20 | 0.72 |
| RVSV, mL | 80 ± 17 | 80 ± 19 | 0.95 |
| RV ejection fraction, % | 56 ± 8 | 56 ± 8 | 0.95 |
| RA area, m^2^ | 21 [18-24] | 20 [18-24] | 0.95 |
| LA volume, mL | 79 [62-97] | 87 [74-102] | 0.20 |
| LGE Type |  |  | 0.42 |
| Nil, n (%) | 15 (42) | 25 (51) |  |
| Non-ischaemic, n (%) | 7 (19) | 5 (10) |  |
| Ischaemic, n (%) | 13 (36) | 19 (39) |  |
| Both, n (%) | 1 (3) | 0 (0) |  |
| ECV fraction, % | 24.9 ± 2.3 | 25.1 ± 1.9 | 0.38 |
| Interstitial volume, mL | 24.1 [21.5-26.9] | 24.0 [20.9-27.9] | 0.72 |
| Myocyte volume, mL | 73.3 [63.4-90.2] | 72.9 [64.5-82.8] | 0.52 |

Values are given as median [interquartile range], mean ± SD or number (percentage).

**Abbreviations**:; ACEi: Angiotensin-converting enzyme inhibitor; ARB: Angiotensin II receptor blocker; CCB: Calcium channel blocker; AHI: Apnoea-hypopnoea index; LVEDV: Left ventricular end-diastolic volume; LVESV: Left ventricular end-systolic volume; LVSV: Left ventricular stroke volume; GLS: Global longitudinal strain; RVEDV: Right ventricular end-diastolic volume; RVESV: Right ventricular end-systolic volume; RVSV: Right ventricular stroke volume; RA: Right atrial; LA: Left atrial; LGE: Late gadolinium enhancement; ECV: Extracellular volume.

**Supplemental Table 2. Univariable linear regression models of clinical, sleep and CMR variables associated with ECV fraction change.**

|  | **Univariable Model** | |
| --- | --- | --- |
|  | **R^2^** | **p Value** |
| **Clinical** |  |  |
| Age, years | 0.002 | 0.72 |
| Male sex | 0.001 | 0.71 |
| ∆Body surface area, m^2^ | 0.000 | 0.85 |
| Obesity (BMI ≥30kg/m^2^) | 0.060 | 0.03 |
| ∆24-hour mean BP, mmHg | 0.007 | 0.47 |
| Diabetes | 0.001 | 0.78 |
| Coronary artery disease | 0.000 | 0.95 |
| Medications, n (%) |  |  |
| Beta Blocker | 0.004 | 0.56 |
| ACEi or ARB | 0.020 | 0.18 |
| CCB | 0.007 | 0.47 |
| Diuretic | 0.002 | 0.67 |
| Statin | 0.012 | 0.32 |
| **Sleep** |  |  |
| Baseline AHI, events per hour | 0.003 | 0.61 |
| Residual (month 12) AHI, events per hour* | 0.003 | 0.71 |
| ∆AHI, events per hour* | 0.007 | 0.26 |
| **CMR Markers** |  |  |
| ∆LVEDV, mL | 0.010 | 0.35 |
| ∆LVESV, mL | 0.003 | 0.60 |
| ∆LVSV, mL | 0.007 | 0.46 |
| ∆LV mass, g | 0.006 | 0.47 |
| ∆LV ejection fraction, % | 0.000 | 0.99 |
| ∆GLS, % | 0.014 | 0.29 |
| ∆RVEDV, mL | 0.011 | 0.35 |
| ∆RVESV, mL | 0.008 | 0.43 |
| ∆RVSV, mL | 0.004 | 0.59 |
| ∆RV ejection fraction, % | 0.004 | 0.58 |
| ∆RA area, m^2^ | 0.014 | 0.28 |
| ∆LA volume, mL | 0.004 | 0.58 |

**Abbreviations**: BMI: Body mass index; ACEi: Angiotensin-converting enzyme inhibitor; ARB: Angiotensin II receptor blocker; CCB: Calcium channel blocker; AHI: Apnoea-hypopnoea index; LVEDV: Left ventricular end-diastolic volume; LVESV: Left ventricular end-systolic volume; LVSV: Left ventricular stroke volume; GLS: Global longitudinal strain; RVEDV: Right ventricular end-diastolic volume; RVESV: Right ventricular end-systolic volume; RVSV: Right ventricular stroke volume; RA: Right atrial; LA: Left atrial.

*Follow-up data only available for CPAP group
